# Meditation practice, mindfulness and pain-related outcomes in mindfulness-based treatment for episodic migraine

**DOI:** 10.1101/2022.01.20.22269474

**Authors:** Carly Hunt, Janelle Letzen, Samuel R Krimmel, Shana A.B. Burrowes, Jennifer A. Haythornthwaite, Michael Keaser, Matthew Reid, Patrick Finan, David A. Seminowicz

**Affiliations:** Johns Hopkins University School of Medicine, Department of Psychiatry and Behavioral Sciences, Baltimore, MD, USA; Boston University School of Medicine, Section of Infectious Diseases, Department of Medicine, Boston MA, USA, 02218; Department of Neurology, Washington University School of Medicine, St. Louis, MO 63110, USA; Department of Neural and Pain Sciences, School of Dentistry, University of Maryland Baltimore, Baltimore, MD, USA 21201; Center to Advance Chronic Pain Research, University of Maryland Baltimore, Baltimore, MD, USA 21201

## Abstract

**Objectives:** Mindfulness-based interventions (MBIs) have emerged as promising prophylactic episodic migraine treatments. The present study investigated biopsychosocial predictors and outcomes associated with formal, daily-life meditation practice in migraine patients undergoing MBI, and whether augmented mindfulness mechanistically underlies change.

**Methods:** Secondary analyses of clinical trial comparing data 12-week mindfulness-based stress reduction (MBSR+; n = 50) to stress management for headache (SMH; n = 48) were conducted.

**Results:** Pre-treatment mesocorticolimbic system functioning (i.e., greater resting state ventromedial prefrontal cortex-right nucleus accumbens [vmPFC-rNAC] functional connectivity) positively predicted meditation practice duration over MBSR+ (*r* = .58, *p* = .001), and moderated change in headache frequency from pre to post-treatment (*b* = -12.60, *p* = .02) such that patients with greater vmPFC-rNAC connectivity showed greater reductions in headache frequency. Patients who meditated more showed greater increases in mindfulness (*b* = .52, *p* = .02) and reductions in the helplessness facet of pain catastrophizing (*b* = -.13, *p* = .01), but not headache frequency, severity or impact. Augmented mindfulness mediated reductions in headache impact resulting from MBSR+, but not headache frequency.

**Conclusions:** Mesocorticolimbic system function is implicated in motivated behavior, and thus could be a target of augmentative interventions designed to enhance meditation practice engagement. Meditation practice appears to benefit pain-related cognitions, but not clinical pain, while mindfulness emerges as a mechanism of MBIs on headache impact, but not frequency. Further research is needed to investigate the day-to-day effects of meditation practice on pain, and continue to characterize the specific mechanisms of MBIs on headache outcomes.

## 1. Introduction

Episodic migraine is a severe and debilitating chronic pain disorder (Leonardi & Raggi, 2013), and nonpharmacological treatments have been established as a key aspect of migraine management (Penzien et al., 2015; Pryse-Phillips et al., 1998). Mindfulness-based stress reduction (MBSR), a group-based skills development program that trains participants in non-judgmental acceptance of experiences (including pain) as they unfold, has recently emerged as a promising nonpharmacological treatment for headache and migraine pain (Gu et al., 2018; Seminowicz et al., 2020; Wells et al., 2020), and related outcomes (e.g., disability; Wells et al., 2014). However, three key aspects of mindfulness-based treatment for headache remain poorly characterized, limiting treatment optimization efforts: 1) biopsychosocial factors that predict participant engagement in the substantial out-of-class, formal meditation practice requirements prescribed in mindfulness-based treatment (henceforth termed “meditation practice”), 2) the effect of meditation practice on clinical pain and psychosocial outcomes, and 3) the extent to which augmented mindfulness, a key putative mechanism of mindfulness-based treatment (Day et al., 2014; Gu et al., 2015) and sequela of meditation practice, mechanistically drives improved outcomes. These three questions, which are critical for the determination of how and for whom mindfulness-based interventions (MBIs) work in migraine, are the foci of the present report.

### Meditation practice in mindfulness-based treatment

Health behaviors like regular meditation practice are difficult to initiate and maintain (Bandura, 2004; King et al., 2009), and little is known about the uptake and impact of meditation practice in the context of MBIs for chronic pain. In healthy samples, personality factors (i.e., openness, conscientiousness) and positive affect may support meditation uptake, while depressive symptoms, perceived stress and neuroticism may interfere (Barrett et al., 2019). In the chronic pain context, the neurobehavioral model of pain, mesocorticolimbic circuitry, and treatment adherence (Letzen et al., 2019) identifies mesocorticolimbic system functioning as a neurobiological mechanism underlying treatment motivation and adherence, and thus a potentially informative predictor of meditation practice engagement. Specifically, the mesolimbic system (i.e., neurons in the ventral tegmental area that project to the nucleus accumbens [NAc], thalamus, amygdala and hippocampus) and its cortical projections (i.e., to the anterior cingulate cortex, ventromedial prefrontal cortex [vmPFC] and orbitofrontal cortex) are activated in the context of approach and avoidance behaviors, respectively (Cardinal et al., 2002; Salamone, 1994; Salamone & Correa, 2012). Individuals with chronic pain show general deficits in mesocorticolimbic functioning and associated reward processing (Loggia et al., 2014) with the degree of reward system dysfunction correlating with pain severity (Kim et al., 2021). Conceivably, patients showing a particularly high degree of mesocorticolimbic system dysfunction may struggle to engage in home meditation practice and thus not benefit from treatment to the same degree as those with lesser dysfunction.

Regarding the effects of meditation practice on pain, the few studies in chronic pain samples addressing this issue suggest that meditation practice associates with improved psychosocial outcomes (e.g., quality of life, psychological distress), but not reduced pain (Pradhan et al., 2007; Rosenzweig et al., 2010). Relatedly, in a large MBSR study of adults with various medical conditions, including chronic pain, meditation practice was associated with favorable changes in mindfulness and other psychological factors, but not medical symptoms (Carmody & Baer, 2008). Outside the pain context, two systematic reviews indicate that the relationship between meditation practice and health-related outcomes is infrequently investigated in MBI studies, and that conclusions are inconsistent on whether the amount of formal practice impacts outcomes (Lloyd et al., 2018; Parsons et al., 2017). It is critical to clarify the link between meditation practice and treatment-induced changes in clinical pain and key psychosocial risk and resilience factors (e.g., mindfulness, pain catastrophizing) to guide recommendations on the optimal meditation practice dosage in MBIs for pain.

### Mindfulness as a treatment mechanism

Theoretically, meditation practice is thought to contribute to increases in mindfulness in the context of MBIs, in turn leading to improved outcomes. Although augmented mindfulness is theorized as a key mechanism of MBIs (Day et al., 2014), for which there is supportive evidence in the non-pain mental health literature according to meta-analytic work (Gu et al., 2015), whether augmented mindfulness mechanistically contributes to pain reductions in headache or other pain conditions remains unknown. Even the direct effect of mindfulness-based treatment on mindfulness in chronic pain appears inconsistent, with MBI efficacy studies variously reporting no significant change in mindfulness in the MBI group or active control (Morone et al., 2009), significant *decreases* in both MBI and active control arms (Dowd et al., 2015), or selective increases in the MBI arm and not active control (Schmidt et al., 2011). It is necessary to investigate whether mindfulness increases in response to mindfulness-based treatment and if such increases underlie pain reductions, to build an understanding of how MBIs work for migraine.

### The present study

Given the above considerations, the present study addressed three aims via secondary analyses of clinical trial data comparing the effects of an enhanced (i.e., 12-week) mindfulness-based stress reduction protocol (MBSR+) versus an active control (stress management for headache; SMH) on clinical pain and neuroimaging outcomes (Burrowes et al., 2021; Seminowicz et al., 2020). First, we investigated the relationship between mesocorticolimbic system functioning, indexed by mesocorticolimbic system functional connectivity (FC) measured pre-treatment, and baseline headache characteristics (frequency, severity and impact), the duration of home meditation practice across MBSR+, and treatment response (i.e., reductions in headache frequency, pain and impact). FC reflects correlated activity among spatially distinct brain regions (Friston, 1994), and has been identified as a useful metric of network level dysfunction in clinical samples (Fox & Greicius, 2010). We hypothesized that greater mesocorticolimbic dysfunction would predict greater baseline pain, reduced meditation practice duration, and lesser treatment gains. We did not specify a priori the direction of FC (i.e., hypoconnectivity versus hyperconnectivity), as both hyper- and hypoconnectivity in mesocorticolimbic circuits have been associated with psychological dysfunction (Fornito & Bullmore, 2015) and been theorized to reflect mesocorticolimbic system dysfunction in chronic pain (Letzen et al., 2019). Second, we investigated the relationship between meditation practice and headache outcomes, mindfulness and pain catastrophizing. Third, we investigated the relationship between dispositional mindfulness and mindfulness-based training, anticipating that MBSR+ would selectively lead to increased mindfulness (relative to SMH) and mechanistically contribute to the improvements in headache frequency and impact that we previously reported in our main clinical trial outcomes paper (Seminowicz et al., 2020).

## 2. Method

The full methodology of the parent clinical trial (R01AT007171) has been described elsewhere (Seminowicz et al., 2020). Here we detail methods pertinent to this secondary analysis.

### 2.1 Participants

Data were drawn from meditation naïve individuals (n = 98) who were randomized to either 12-week MBSR+ (n = 50) or SMH (n = 48). Participants were recruited from 2014 to 2017. Participants were included if they met criteria for the International Classification of Headache Disorders criteria for migraine without aura and had been living with migraine for at least 1 year. Participants were excluded if they reported severe psychiatric symptoms, opioid medication use, prior mindfulness experience, or engagement in any treatment anticipated to impact mindfulness training (see the parent project Protocol for full inclusion and exclusion criteria). Demographic characteristics are shown in Table 1.

**Table 1.** Sample demographic and baseline clinical pain characteristics

|  | MBSR+ | SMH |
| --- | --- | --- |
| Age, mean (SD) | 39.1 (12.7) | 37.2(12.3) |
| Gender, n (%) |  |  |
| M | 3 (6.0) | 6 (12.5) |
| F | 47 (94.0) | 42 (87.5) |
| Race, n (%) |  |  |
| White | 35 (71.4) | 36 (75) |
| Black | 10 (20.4) | 7 (14.6) |
| Other | 4 (8.0) | 5 (10.4) |
| College Graduate, n (%) | 37 (74.0) | 41 (85.4) |
| Headache Frequency, mean (SD) | 8.42 (2.51) | 8.42 (3.3) |
| Headache Severity, mean (SD) | 4.7 (1.7) | 4.3 (1.6) |
| Headache Impact, mean (SD) | 60.1 (5.8) | 60.9 (5.6) |
| Average Meditation Practice Duration | 20.8 (10.0) | - |

*Notes.* MBSR+ = Enhanced Mindfulness-Based Stress Reduction; SMH = Stress Management for Headache. Average meditation practice duration reflects minutes meditated per day on average over the course of MBSR+.

### 2.2 Interventions

Participants were randomized to either MBSR+ or SMH according to procedures previously described (Seminowicz et al., 2020). Each condition included 12, 2-hour classes over 4 months, including 8 weekly sessions followed by 4 bi-weekly sessions. MBSR+ was adapted from Jon Kabat Zinn’s MBSR program, and included a 4-hour retreat between week 6 and 8 in addition to the 2-hour classes. MBSR+ participants were given audio CDs and handouts to support home practice, as well as a copy of *Full Catastrophe Living* by Jon Kabat-Zinn. The 4 classes that were held beyond the typical 8-week MBSR program focused on encouraging continued practice of mindfulness, as well as on positive psychological constructs often emphasized in Buddhist-derived meditation training (e.g., equanimity and self-compassion). SMH included didactic content on the effect of stress and other triggers in migraine, and was similar in format to MBSR+ except that it did not include a retreat or homework. Topics included stress and coping, sleep hygiene, pain education, and migraine medications. Giving and receiving social support, group discussion and information were emphasized, whereas skill development and behavior change were not covered. Each session included 10 minutes of standardized muscle stretching practices, and participants were given a copy of *The Migraine Brain* by Carolyn Bernstein.

### 2.3 Measures

Demographic and neuroimaging data were collected at baseline. Clinical pain and psychosocial variables were collected at baseline, at 10 weeks (after the completion of 8 sessions), at 20-weeks (after 12 sessions) and at week 52, although mindfulness was not measured at week 52.

#### 2.3.1 Headache Frequency, Severity and Impact

Headache frequency was measured using a 28-day electronic diary based on the National Institute of Neurological Disorders and Stroke preventive therapy headache diary, which participants accessed through an emailed link. In cases where fewer than the full 28 days were completed, the proportion of headache days was calculated (number of headache days/total number of diary days) and then multiplied by 28 to get a continuous headache days variable. Headache severity was assessed with a 0-10 scale, and was quantified as the average of all headache intensity ratings from the diary. Headache impact was measured with the Headache Impact Test (HIT-6), which assesses the effects of headaches on lifestyle (Kosinski et al., 2003); it has been validated in chronic and episodic migraine (Yang et al., 2011).

#### 2.3.2 Psychosocial Constructs and Meditation Practice Duration

Mindfulness was measured with the Five Factor Mindfulness Questionnaire (FFMQ; Baer et al., 2006), which yields 5 subscale scores and a total score. Participants rate items on 5-point Likert scales (1 = *never or very rarely true*; 5 = *very often or always true*). Total scores were computed by taking the sum of each subscale score, and were used in analyses. Pain catastrophizing was measured with the pain catastrophizing scale (PCS; Sullivan et al., 1995), a 13-item measure which asks participants to rate how often they experience particular feelings and thoughts when experiencing pain (0 = *not at all*, 4 = *all the time*). The PCS contains 3 subscales assessing specific aspects of pain catastrophizing, 1) rumination (e.g., “I can’t stop thinking about how much it hurts”, 2) magnification (“I’m afraid that something serious might happen”), and 3) helplessness (“There is nothing I can do to reduce the intensity of my pain”), as well as a total score. Consistent with prior studies (Parsons et al., 2017), participants completed weekly paper home practice logs where they reported minutes of formal meditative practice completed per day (e.g., sitting meditation, body scan, yoga) each day of that week, and submitted the log during that week’s class. The amount of meditation practice was quantified as the average number of minutes spent in meditation per day over the course of MBSR+.

#### 2.3.3 Mesocorticolimbic System Functional Connectivity

Functional connectivity of the mesocorticolimbic system was probed using nucleus accumbens (NAc) regions of interest (ROIs) in whole-brain, seed-based functional connectivity analyses. Details on neuroimaging data acquisition, processing, and first-level analyses are described in Supplemental Material.

#### 2.3.4. Group-Level Resting-State Functional Connectivity Analyses

Individuals’ first-level contrast maps were entered into a group-level analysis. Across the sample, mean time practicing meditation was used as a regressor of interest in probing functional connectivity between each NAc seed and the whole brain. Data were examined at the group-level using Random Field Theory (Worsley et al., 1996) parametric statistics with a combination of an uncorrected height threshold (*p* < .001) and false discovery rate-corrected cluster-level threshold (*p*FDR < .05) as is standard criterion in the CONN toolbox. *A priori*, we planned that individual-level values from significant clusters emerging from this analysis would be extracted and included in subsequent analyses with behavioral data to further understand aspects of adherence to meditation practice.

### 2.7 Statistical Approach

Means and standard deviations were calculated for demographic variables, primary measures and covariates. Outcome variables were inspected for normality and outliers (+/- 3 *SD* around the mean), and outliers were removed. The relationship between mesocorticolimbic system functioning, baseline clinical pain and meditation practice during MBSR+ were quantified with bivariate Pearson’s correlations. Moderation hypotheses (i.e., the moderation effect of mesocorticolimbic system functioning on change in clinical pain, and the moderation effect of meditation practice duration on change in clinical pain and psychosocial outcomes) were tested using linear mixed models (Singer & Willett, 2003) allowing for random intercepts. Dummy codes were created for follow up time points (i.e., mid-treatment [10 weeks], post-treatment [20 weeks], and baseline to 1-year follow up [52 weeks]), with baseline entered as the reference group. Consistent with the analytic approach taken for the primary clinical trial data (Seminowicz et al., 2020), pre to post-treatment comparisons were considered primary, and other comparisons secondary. Linear mixed models were specified and visualized using the *lme4, lmerTest, interactions* and *jtools* R packages (Bates et al., 2015; Kuznetsova et al., 2017; Long, 2019, 2020) using restricted maximum likelihood estimation. Significance values were calculated using Satterthwaite’s method (see Kuznetsova et al., 2017). We report the marginal R^2^ for linear mixed models, which is the outcome variance explained by the fixed effects, as well as the conditional R^2^, which is the variance explained by both fixed and random effects (Nakagawa et al., 2017).

Mediation analyses were specified in mPlus (Muthen & Muthen, 1998-2017) in accordance with recommendations for two-wave longitudinal mediation models (MacKinnon, 1994) using maximum likelihood estimation. Confidence intervals were estimated using bias-corrected bootstrapping and used to interpret the significance of mediated effects (MacKinnon et al., 2004). Model fit was evaluated using the chi square test of model fit, Root Mean Square Error of Approximation (RMSEA), Comparative Fit Index (CFI), and Standardized Root Mean Square Residual (SRMR) and recommended cutoff scores (Hu & Bentler, 2009). Specifically, in path models, we tested whether treatment group assignment led to changes in post-treatment mindfulness, which in turn led to changes in clinical pain, adjusting for baseline clinical pain and mindfulness.

## 3. Results

### 3.1 Descriptive Data

Forty-seven of the 50 participants randomized to the MBSR+ condition provided some amount of home practice information. Across those 47 participants, the meditation diary completion rate was 95% (i.e., there were 3066 reportable days (among participants who attended the session and turned in the diary), and participants provided meditation duration information on 2916 of these days). Across visits, participants meditated on average 20.8 minutes per day (*SD* = 10.0; range = 3.78 – 46.75). As described in the clinical trial primary outcomes paper (Seminowicz et al., 2020), just 3 participants across MBSR+ and SMH were lost to follow-up. Nearly all participants in the MBSR+ group (48 or 49 out of the 50 randomized) provided clinical pain, pain catastrophizing and mindfulness data at 12-weeks.

### 3.1 Mesocorticolimbic functional connectivity

Using the left NAc (lNAc) and right NAc (rNAC) ROIs as seeds with no other regressors in the model, we observed significant associations within clusters spanning key regions of the mesocorticolimbic system, including bilateral medial prefrontal cortex, orbitofrontal cortex, ventral tegmental area, amygdala, and hippocampus (Table S1 for a full list of regions; Figure 1) among MBSR+ participants. When average meditation practice duration was added to the model as a predictor of seed-to-voxel functional connectivity, the rNAc seed continued to show significant functional connectivity with a cluster comprising the vmPFC (Table 2; Figure 2). However, the lNAc seed did not show any significant functional connectivity patterns. In other words, meditation practice predicted significant seed-to-voxel functional connectivity between rNAc and vmPFC but did not predict lNAc functional

**Fig 1.**
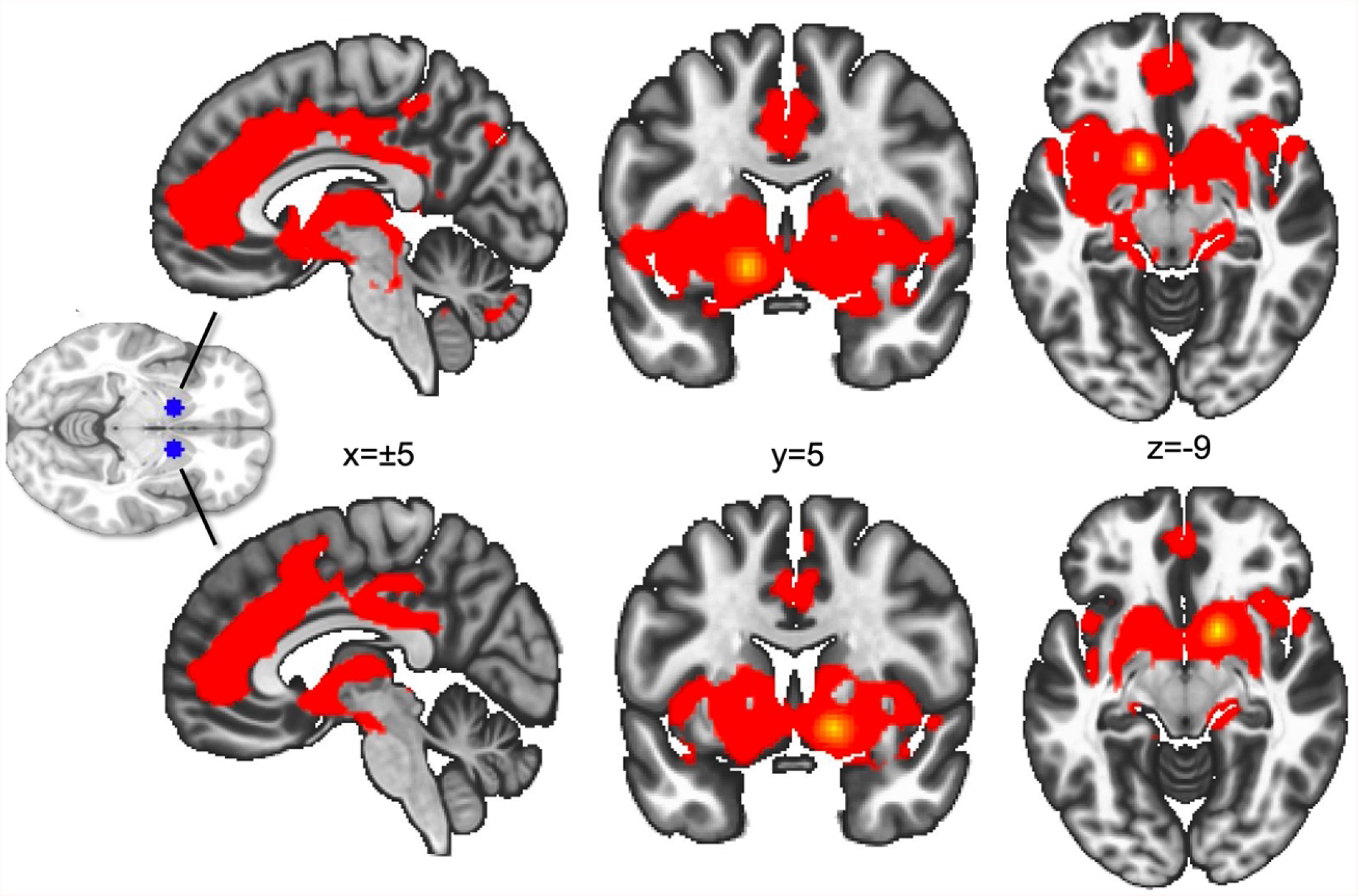
Whole-brain functional connectivity based on left (top) and right (bottom) nucleus accumbens regions of interest (ROIs). ROIs used as seeds are shown in blue

**Fig 2.**
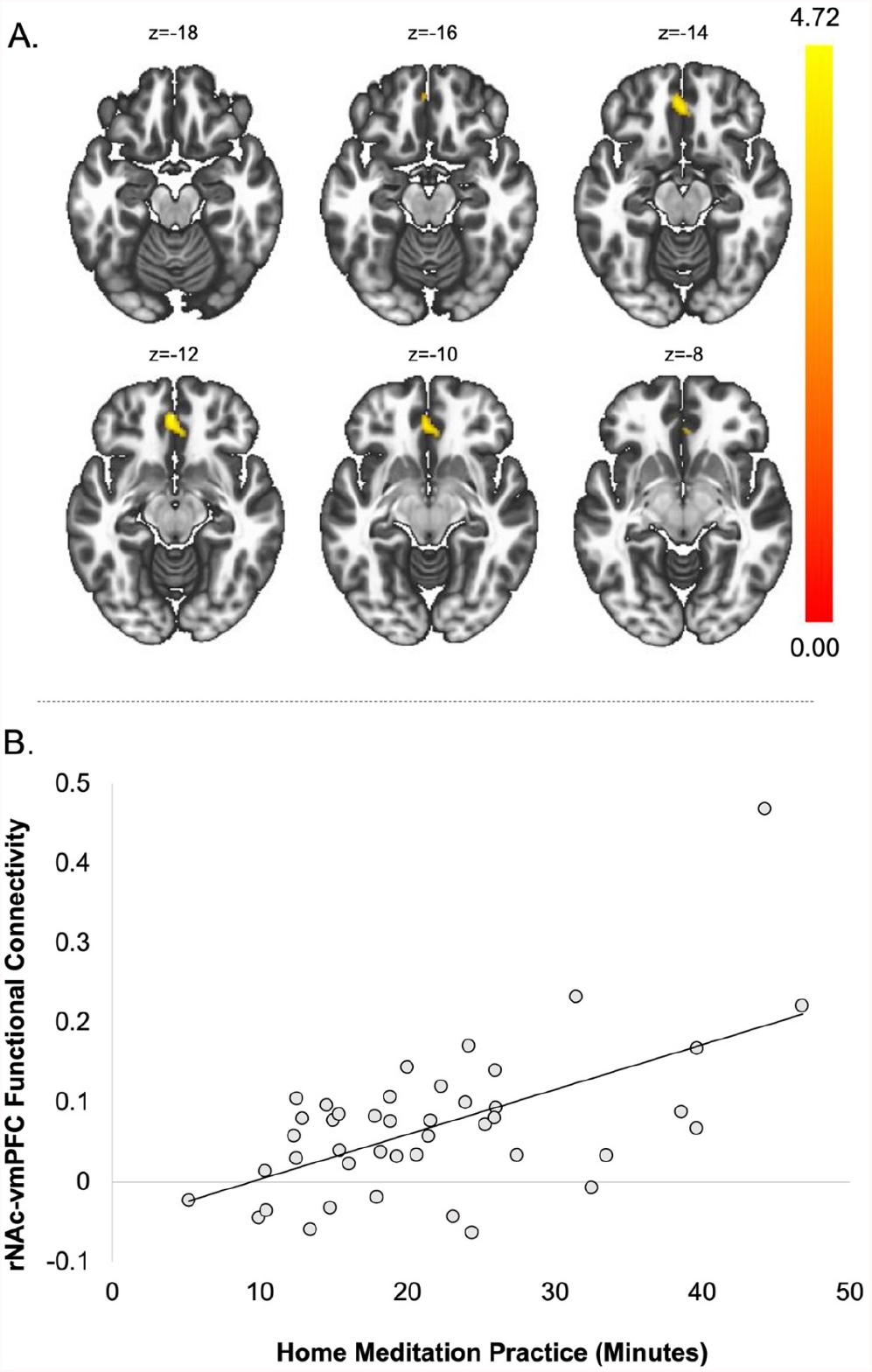
*A*. Using meditation practice as a predictor, there was significant functional connectivity between the right nucleus accumbens (rNAc) seed and a cluster spanning the ventromedial prefontal cortex (vmPFC) after multiple comparisons corrections. *B*. Association between rNAC-vmPFC functional connectivity values and meditation practice

**Table 2.**
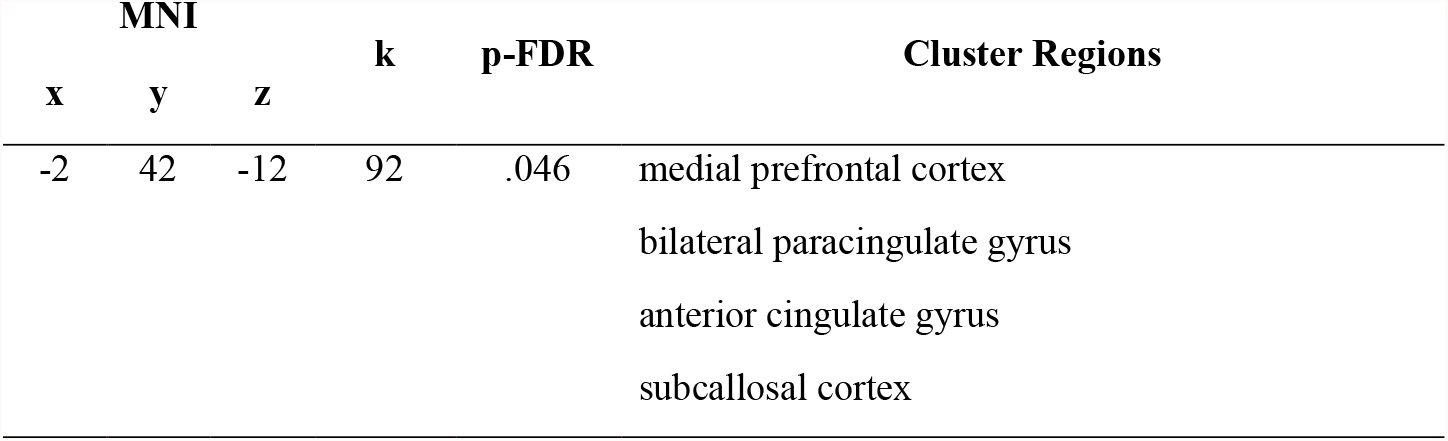
At-home meditation practice as a predictor of whole-brain functional connectivity using the right nucleus accumbens seed

### 3.2 Mesocorticolimbic functioning, meditation practice and headache outcomes

Individual-level values from the significant rNAc-vmPFC cluster were extracted and entered in statistical models with behavioral data. rNAC-vmPFC FC was positively correlated with average meditation practice duration over the course of the intervention (*r* = .58, *p* = .001). Means and standard deviations for baseline headache frequency, severity and impact are shown in Table 1. rNAC-vmPFC connectivity was unassociated with baseline headache frequency (*r* = .09, *p* = .57), severity (*r* = .08, *p* = .63), and impact (*r* = -.06, *p* = .69).

As we reported previously, a significant group by time (post-treatment versus baseline) interactions were observed for HA frequency and impact such that MBSR+ participants exhibited greater declines across treatment (Seminowicz et al., 2020). Among MBSR+ participants, adherence-associated rNAc-vmPFC connectivity significantly moderated the change in headache frequency from baseline to post-treatment (*b* = -12.60, se = 5.36, *p* = .02, model R^2^ _marginal_ = .19, R^2^ _conditional_ = .54), such that greater declines in headache frequency were observed among participants with greater rNAC-vmPFC connectivity (Figure 3). rNAC-vmPFC connectivity did not moderate the change in average headache pain severity or headache impact from baseline to post-treatment (*p*’s > 05; see Table S2). Adherence-associated rNAC-vmPFC connectivity did not moderate change in headache frequency from baseline to mid-treatment (*b* = -8.41, se = 5.36, *p* = .12), or baseline to 52-weeks (*b* = -7.2, se = 5.29, *p* = .18), or change in headache pain severity or impact from baseline to mid-treatment, and baseline to 1-year follow-up (*p*’s > .05; full statistics available in Table S2).

**Fig 3.**
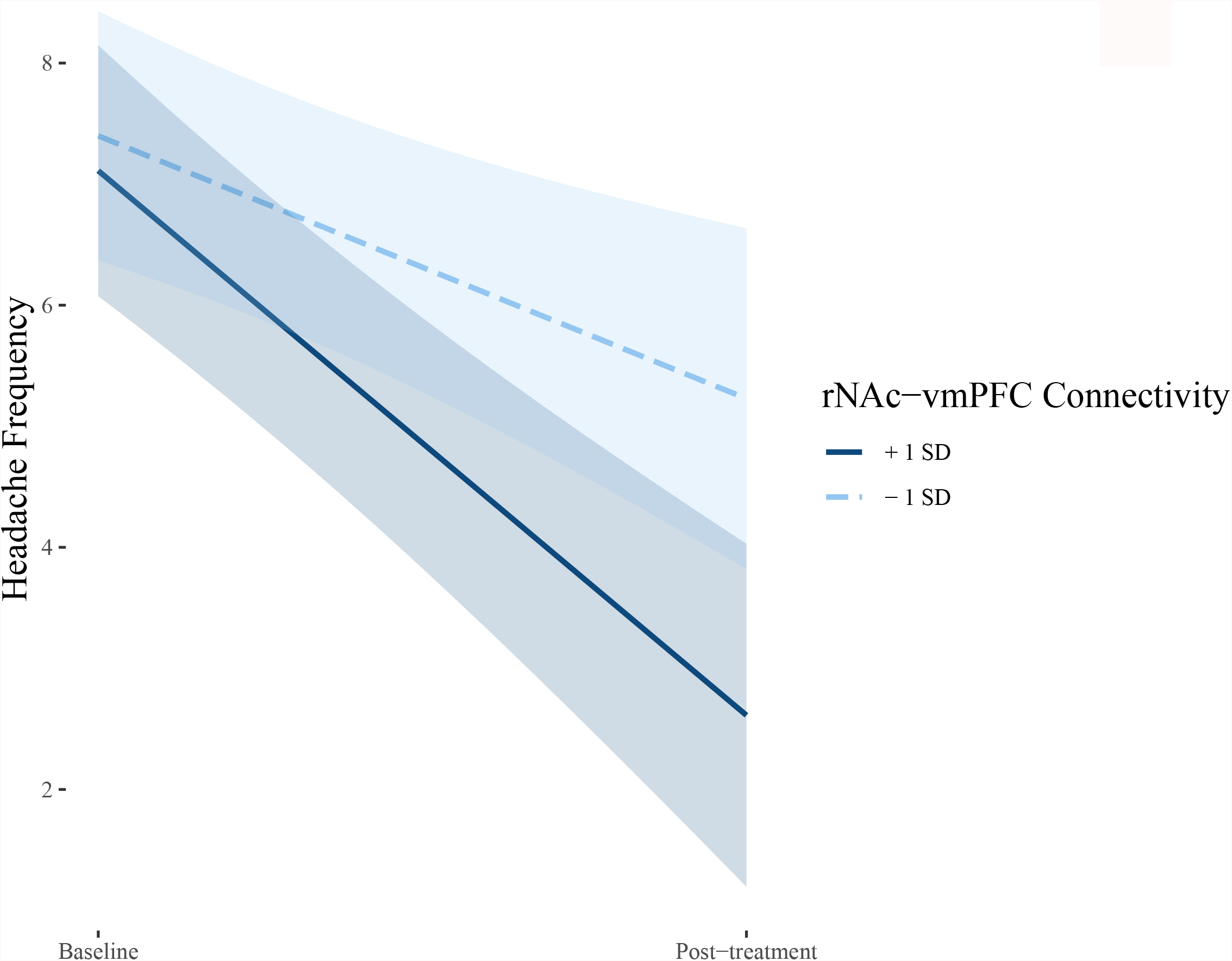
Participants in the MBSR+ demonstrating higher levels of rNAc-vmPFC connectivity showed greater reductions in headache frequency from pre to post treatment

### 3.3 Effects of meditation practice on mindfulness, headache outcomes, and pain reactivity

Meditation practice did not moderate change in headache frequency (*b* = -.07, *se* = .05, *p* = .14), severity (*b* = .01, *se* = .02, *p* = .71), or impact (*b* = -.002, *se* = .09, *p* = .98), from baseline to 20-weeks or from baseline to mid-treatment or 1-year follow up (*p*’s > .05, full statistics shown in Table S3). Meditation practice moderated change in pain catastrophizing-helplessness from baseline to 20 weeks (*p* = .01; Table 3; Figure 4A), such that those who meditated more appeared to start higher in helplessness, and show greater reductions from pre to post-treatment. Meditation practice moderated the change in mindfulness from baseline to 20 weeks (*p* = .02; Figure 4B), such that individuals who meditated more saw greater gains in mindfulness. Meditation practice did not moderate change in pain catastrophizing-helplessness from baseline to mid-treatment or 1-year follow up, and had no moderating effect on change in magnification, rumination, or total pain catastrophizing scores at any time point (*p*’s > .05; see Table 3). Meditation practice did not moderate change in mid-treatment mindfulness (*p* > .05; Table 3), and mindfulness was not measured at 1-year follow-up.

**Table 3.** Relationship between meditation practice and change in pain catastrophizing and mindfulness over the course of MBSR+

| *Predictors* | **PCS Total** |  |  | **PCS Helplessness** |  |  | **PCS Magnification** |  |  | **PCS Rumination** |  |  | **Trait Mindfulness** |  |  |
| --- | --- | --- | --- | --- | --- | --- | --- | --- | --- | --- | --- | --- | --- | --- | --- |
|  | *Estimates* | *SE* | *p* | *Estimates* | *SE* | *p* | *Estimates* | *SE* | *p* | *Estimates* | *SE* | *p* | *Estimates* | *SE* | *p* |
| Intercept | 8.58 | 2.48 | **<0.001** | 1.71 | 1.14 | 0.13 | 1.25 | 0.48 | **0.01** | 5.01 | 1.22 | **<0.001** | 139.88 | 6.43 | **<0.001** |
| Mid-Treatment | 0.50 | 2.46 | 0.84 | 1.08 | 1.21 | 0.37 | 0.54 | 0.53 | 0.31 | -0.51 | 1.31 | 0.70 | 1.65 | 5.17 | 0.75 |
| Home Practice | 0.09 | 0.11 | 0.39 | 0.11 | 0.05 | **0.03** | 0.02 | 0.02 | 0.29 | -0.01 | 0.05 | 0.89 | 0.08 | 0.28 | 0.76 |
| Post-Treatment | 0.50 | 2.46 | 0.84 | 1.36 | 1.21 | 0.26 | -0.73 | 0.54 | 0.17 | 0.49 | 1.30 | 0.70 | -1.36 | 5.17 | 0.79 |
| 1-year Follow-Up | 0.22 | 2.57 | 0.93 | 0.83 | 1.26 | 0.51 | 0.30 | 0.56 | 0.60 | -0.59 | 1.35 | 0.66 |  |  |  |
| Mid-Treatment X<br>Meditation Practice | -0.10 | 0.11 | 0.36 | -0.08 | 0.05 | 0.12 | -0.04 | 0.02 | 0.11 | -0.01 | 0.06 | 0.83 | 0.03 | 0.22 | 0.90 |
| Post-Treatment X<br>Meditation Practice | -0.18 | 0.11 | 0.08 | -0.13 | 0.05 | **0.01** | -0.00 | 0.02 | 0.99 | -0.08 | 0.06 | 0.14 | 0.52 | 0.22 | **0.02** |
| 1-Year Follow-up X<br>Meditation Practice | -0.00 | 0.11 | 0.97 | -0.02 | 0.06 | 0.67 | -0.02 | 0.02 | 0.41 | 0.01 | 0.06 | 0.83 |  |  |  |
| **Random Effects** |  |  |  |  |  |  |  |  |  |  |  |  |  |  |  |
| $\sigma^2$ | 25.37 | | | 6.17 | | | 1.16 | | | 7.08 | | | 104.56 | | |
| $\tau_{00}$ | 28.00<sub>id</sub> | | | 5.13<sub>id</sub> | | | 0.76<sub>id</sub> | | | 5.86<sub>id</sub> | | | 239.88<sub>id</sub> | | |
| N | 47<sub>id</sub> |  |  | 47<sub>id</sub> |  |  | 47<sub>id</sub> |  |  | 47<sub>id</sub> |  |  | 47<sub>id</sub> |  |  |
| Observations | 180 |  |  | 180 |  |  | 176 |  |  | 181 |  |  | 137 |  |  |
| Marginal R<sup>2</sup> /<br>Conditional R<sup>2</sup> | 0.046 / 0.546 |  |  | 0.078 / 0.497 |  |  | 0.053 / 0.427 |  |  | 0.032 / 0.470 |  |  | 0.077 / 0.720 |  |  |

Notes. PCS = Pain Catastrophizing Scale. SE = Standard Error. Follow-up time variables (i.e., Mid-Treatment, Post-Treatment and 1-year Follow-Up) were entered as dummy codes, with baseline as the reference group. $\sigma^2$ = L1 variance; $\tau_{00}$ = intercept variance.

**Fig 4.**
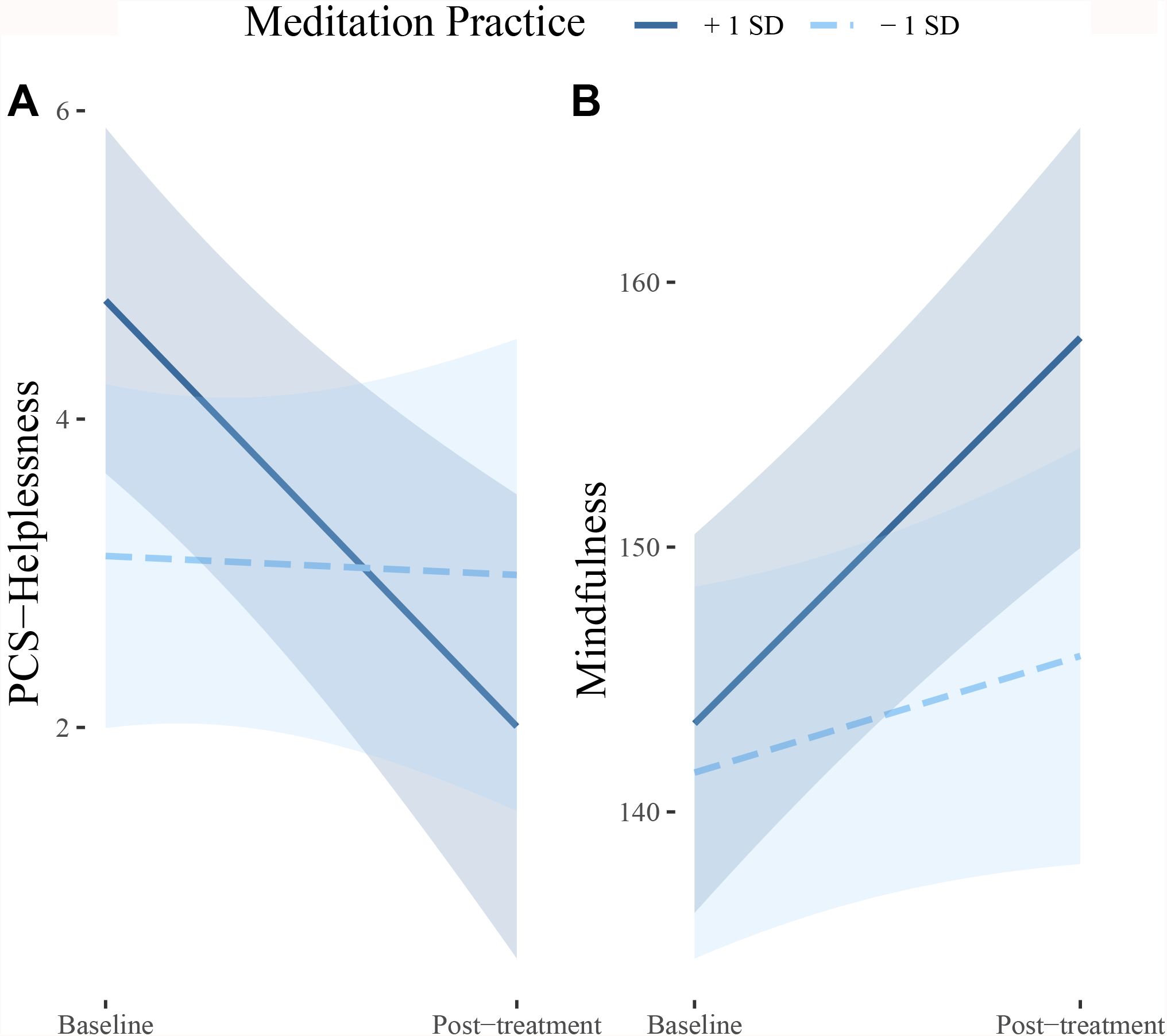
Participants who meditated more on average showed more favorable changes from pre to post-treatment in the helplessness facet of pain catastrophizing and in mindfulness

### 3.4 Augmented mindfulness as a treatment mechanism

Treatment group significantly moderated the change in mindfulness from baseline to post-treatment (*b* = 7.22, *se* = 1.50, *t* = 4.82, *p* < .001; model R^2^ _marginal_ = 0.05, R^2^ _conditional_ = .73), such that mindfulness increased significantly in the MBSR+ group (*b* = 10.65, *se* = 2.05, *t* = 5.20, *p* < .001), but not in SMH (*b* = 3.52, *se* = 2.12, *t* = 1.67, *p* = .10; Figure S1). As described in Methods, we investigated whether the treatment-induced change in mindfulness mediated the significant between-groups differences in headache frequency and headache impact that we reported in our clinical trial outcomes paper (Seminowicz et al., 2020). The mediation model of headache impact fit the data well as indicated by most fit indices, including the chi-square test of model fit (chi square = 6.5, *df* = 2, *p* = .04), CFI (.95) and SRMR (.05), although the RMSEA equaled .16 (probability RMSEA <= .05 = .07). Confidence intervals for the indirect effect of treatment group assignment on headache impact via mindfulness indicated a significant indirect effect (point estimate = -.57, 95% CI: -0.10,-.01), such that MBSR+ led to greater post-treatment mindfulness adjusting for baseline mindfulness relative to SMH (*b* = 7.85, *p* = .008), which in turn led to reduced post-treatment headache impact adjusting for baseline headache impact (b = - .07, p = .010); full results shown in Figure 5. For the model of headache frequency, three fit indices suggested good fit (CFI = 1.00, SRMR = .02, RMSEA = .000), and two suggested inadequate fit (CFI = .92, chi square = .02, df = 2, *p* = .63). The hypothesized indirect effect of treatment on headache frequency via mindfulness was not supported (point estimate = .05, 95% CI: -.24, 4.4).

**Fig 5.**
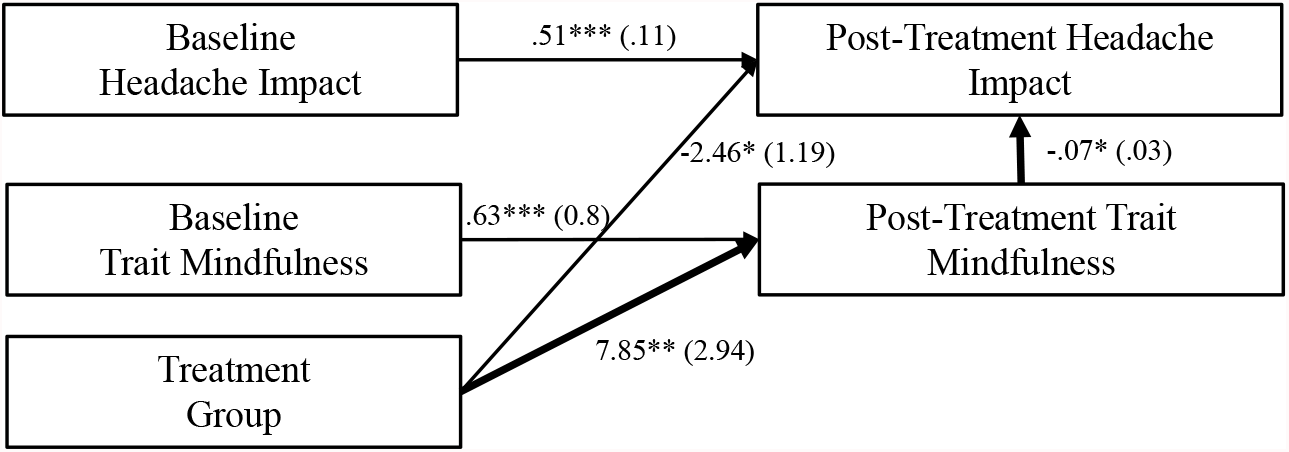
Model of change in trait mindfulness as a mediator of the association between treatment group assignment (MBSR+ versus SMH) and change in headache impact from baseline to post-treatment. Confidence intervals obtained via bias-corrected bootstrapping suggested a significant indirect effect of treatment on headache impact via augmented trait mindfulness (dark bolded path). Unstandardized estimates are shown with standard errors in parentheses. Exogenous variables were allowed to covary. \**p* < .05, \*\**p* < .01, \*\*\**p* < .001

## 4. Discussion

### Mesocorticolimbic functioning, meditation practice and pain outcomes

Patients demonstrating greater connectivity between the rNAc and vmPFC meditated for greater amounts of time each day on average and showed greater reduction in headache frequency over the course of MBSR+, relative to those with lesser connectivity. Given that the NAc and prefrontal cortex (including the vmPFC) have been strongly implicated in motivated behavior (Cardinal et al., 2002; Di Domenico & Ryan, 2017; Knutson et al., 2001), demonstrating augmented intrinsic connectivity during reward motivation (Ballard et al., 2011), these findings suggest that motivational factors may contribute to patient engagement in meditation practice during mindfulness-based treatment. Further, they provide support for the neurobehavioral model of pain, mesocorticolimbic circuitry, and treatment adherence (Letzen et al., 2019), in that mesocorticolimbic functioning significantly predicted treatment adherence and outcomes. Notably, both functional hypoconnectivity and hyperconnectivity have been theorized to indicate mesocorticolimbic reward system dysfunction in chronic pain (Letzen et al., 2019), which is supported by data suggesting that major depression, which like chronic pain is characterized by anhedonia and amotivation, is characterized by patterns of both hyper- and hypoconnectivity (Kaiser et al., 2015). The relative hypoconnectivity of the vmPFC and NAc observed among patients who meditated less represents a relative loss of communication among these regions in comparison with those who showed greater adherence, and may be one neurobiological mechanism underlying low treatment adherence.

The NAc-vmPFC system is involved in the evaluative aspects of pain (Woo et al., 2015) and pain chronification (Baliki et al., 2012), while chronic pain patients demonstrate reduced connectivity between the NAc and frontal regions compared with healthy controls, which associates with risk and reward processing difficulties (Berger et al., 2014). Clinical depression, also characterized by reward processing difficulties, is marked by attenuated NAc-vmPFC connectivity compared with healthy controls (Furman et al., 2011), with the degree of attenuation correlating with symptom severity (Satterthwaite et al., 2015). Although speculative, NAc-vmPFC hypoconnectivity observed in this study could reflect a deficit in positive reward anticipation or processing, in that individuals showing hypoconnectivity anticipated or experienced less reward from meditation practice. Future work should continue to examine the relation between frontostriatal connectivity and treatment adherence in chronic pain. Augmentative therapies targeting mesocorticolimbic functioning and motivational factors, as well as other biopsychosocial factors tied with treatment adherence (e.g., self-efficacy) (Bandura, 2004) could potentially promote greater use and benefit from meditation practice and other pain-self management techniques.

### Effects of meditation practice on mindfulness, clinical pain, and pain reactivity

Meditation practice moderated changes in mindfulness and the helplessness facet of pain catastrophizing, such that those who meditated more saw more favorable improvements in these outcomes. These findings accord with others in non-pain samples showing that meditation practice (Carmody & Baer, 2008) and gains in state mindfulness achieved through meditation (Kiken et al., 2015) promote improved mindfulness. We did not locate other literature describing associations between meditation practice and pain catastrophizing in chronic pain patients; although, formal in-person training has been shown to reduce pain catastrophizing relative to active control conditions (Turner et al., 2016). Perhaps patients who meditated more gained a greater sense of control over their pain (i.e., reduced helplessness), relative to those who practiced less. However, meditation practice did not moderate changes in headache outcomes, suggesting that it favorably modulates psychosocial outcomes but not headache frequency or intensity, consistent with prior MBI studies in other pain conditions (Pradhan et al., 2007; Rosenzweig et al., 2010).

### Mindfulness as a treatment mechanism

Lastly, we investigated the extent to which mindfulness changes in response to MBSR+ and SMH, and mediates the reductions in headache frequency and impact following MBSR+ relative to SMH reported in our main clinical trial outcomes paper (Seminowicz et al., 2020). Consistent with hypotheses, we observed that MBSR+ led to increased mindfulness whereas SMH did not, and augmented mindfulness was a significant mechanism linking MBSR+ practice and headache impact. However, mindfulness did not underlie changes in headache frequency. As such, augmented mindfulness appears to support the benefits of mindfulness-based treatment in terms of the impact of headache on patients’ lives, but not how often headaches occur. This is consistent with theoretical perspectives on how mindfulness training works, as discussed in a recent narrative review of MBIs for migraine (Wells et al., 2020): it is meant to change the patient’s relationship to pain (i.e., through greater non-judgmental acceptance), and not necessarily pain frequency. Besides mindfulness, there could have been other unmeasured MBI-specific psychosocial mechanisms that accounted for the greater improvements in headache frequency at post-treatment in the MBSR+ group relative to SMH; for instance, emotion regulation (Chambers et al., 2009). Notably, our findings are consistent with others showing that mindfulness training uniquely enhances mindfulness relative to control conditions (Schmidt et al., 2011), and inconsistent with others suggesting no unique effect (Anheyer et al., 2019; Morone et al., 2009). The MBI we employed was more intensive than the standardized 8-week MBSR protocol (Kabat-Zinn, 1982), which may have contributed to its capacity to enhance mindfulness to a greater degree than our active control.

### Limitations, conclusions and future directions

In conclusion, findings suggest that mesocorticolimbic functioning predicts greater meditation practice during mindfulness-based treatment for chronic migraine, as well as treatment response. Greater engagement in meditation practice appears to benefit mindfulness and pain catastrophizing, but not headache outcomes, at least during a 12-week intervention and 1-year follow-up period. Augmented mindfulness emerged as a mechanism underlying improvements in headache impact following mindfulness-based training, but not headache frequency. Future directions include replication of the relationship between rNAc-vmPFC connectivity and treatment adherence to mindfulness-based and other psychosocial treatments in chronic pain samples. More broadly, future studies should investigate whether augmentative interventions (e.g., motivational interviewing) can be effectively combined with MBSR to enhance meditation practice engagement. Future research should also employ intensive longitudinal methods (Shiffman & Stone, 1998) to achieve a more granular understanding of the effects of meditation practice on pain and related outcomes on a day-to-day basis, as well as barriers to meditation use, in daily life. Lastly, future research should continue to characterize the mechanisms through which MBIs impact pain.

Several limitations should be noted. We were unable to obtain meditation practice data when participants did not attend the weekly class to turn in their home practice logs, and we lack an objective measure of meditation use (e.g., smartphone meditation app utilization), both of which may have introduced bias. Real-time, smartphone-based assessment of meditation use might be employed in future studies to address these issues. Second, we cannot disentangle whether participants were practicing mindfulness meditation, lovingkindness, or hatha yoga, all of which are taught in MBSR. Third, our results describing the associations between meditation practice and outcomes are simply correlational. Lastly, the study sample was predominately white and well-educated and additional studies are needed to investigate these phenomena in diverse samples. Despite its limitations, this study provides 1) a novel characterization of the link between mesocorticolimbic system function, mindfulness-based treatment adherence and response, 2) further support for the positive relationship between meditation practice and improved psychosocial outcomes in chronic pain, and 3) evidence that mindfulness may indeed be one mechanism of mindfulness-based treatment for migraine.

## Supporting information

STROBE Checklist

## Data Availability

All data produced in the present study are available upon reasonable request to the authors.

## Acknowledgments

This work was supported by National Institutes of Health awards T32NS070201 (CAH) and R01AT007176 (DAS).

