## Supplemental Material for "Meditation practice, mindfulness and pain-related outcomes in mindfulness-based treatment for episodic migraine"

**Neuroimaging methods: Data acquisition, processing, and first-level analyses**

***MRI Acquisition.*** Neuroimaging data were collected using a Siemens 3T Tim Trio scanner with a 32-channel head coil or a Siemens 3T Prisma scanner with a 64-channel head coil at the University of Maryland, Baltimore Medical Imaging Facility. This difference in scanner was due to a scanner upgrade during data acquisition; however, thorough processing of our data showed that this upgrade did not substantially impact data. The relevant scans from the imaging protocol that were used for this study included a T1-weighted structural 3D MPRAGE scan (whole brain coverage, TR = 2300 ms, TE=2.98 ms, voxels = 1.00 mm isotropic) and an eyes-open, resting-state EPI scan while participants fixated on a crosshair (whole brain coverage, TR = 2000 ms, TE = 28 ms, voxels 3.4 × 3.4 × 4.0 mm, slices = 40, duration = 300 TR).

***ROI Selection****.* To examine functional connectivity of the mesocorticolimbic system, we created two lateralized NAc ROIs using the Wake Forest University Pickatlas toolbox (Maldjian et al., 2003, 2004). The NAc is a region within the mesolimbic system that is considered a critical structure for reward processing. Primary afferent fibers to the NAc shell functionally and structurally connect the mesolimbic system with corticolimbic regions, such as the medial prefrontal cortex (Bayassi-Jakowicka et al., 2021). Previous studies have used the NAc as a seed ROI to probe mesocorticolimbic system functional connectivity during resting-state fMRI (Cauda et al., 2011; H. Gu et al., 2010; Zhang et al., 2019).

In the present study, NAc ROIs were constructed by projecting 6-mm spheres around coordinates (±16, 10, -8) used in seminal work that reported on differences in NAc-related function during experimental pain onset and offset (as relief) based on chronic pain status (Baliki et al., 2010). When input in a large-scale, automated synthesis of fMRI data via the Neurosynth platform (Yarkoni et al., 2011), these coordinates yield z-scores of 21.99 (left) and 23.64 (right) for their association with the term “reward” and 17.24 (left) and 14.89 (right) for their association with the term “nucleus accumbens”.

***MRI Data Processing and Denoising.*** SPM12 was used to complete preprocessing. Steps included slice timing correction, realignment (motion correction), coregistration of the T1 to the mean functional image, segmentation of the T1, normalization of functional images with interpolation to 2 × 2 × 2-mm voxels, and smoothing with a 6-mm full width at half maximum (FWHM) Gaussian kernel. Custom scripts were used to conduct motion regression. Participants with FrameWise Displacement Arithmetic Mean greater than 0.3 were removed (Power et al., 2012, 2014, 2015).

Following preprocessing, data were entered in CONN toolbox (version 17f; <http://www.nitrc.org/projects/conn>) for denoising, first-level, and second-level analyses. Global signal was not removed based on ongoing debate (Murphy and Fox, 2017). White matter (WM) and cerebral spinal fluid (CSF) confounding signals were controlled for using the aCompCor algorithm (Behzadi et al., 2007; Muschelli et al., 2014). Because we did not remove global signal, we used eroded WM and CSF masks that did not include external or extreme capsules (Power et al., 2017). Additional denoising steps included the removal of realignment parameters along with their first-order derivatives, simultaneous bandpass filtering (0.008 and 0.09 Hz), linear detrending, and despiking after regression steps (Patel et al., 2014). We visually inspected denoised data to ensure that processing resulted in a normalized distribution of connectivity values for each participant.

***First-Level Analyses.*** The visually inspected, denoised data then underwent first-level analyses in which averaged timeseries data from all voxels within each NAc seed ROI was extracted. Fisher-transformed bivariate correlation coefficients were computed between the NAc ROI’s timeseries and the timeseries of each individual voxel.

*Table S1.* Whole-brain, seed-to-voxel functional connectivity analyses using left and right nucleus accumbens regions of interest (±16, 10, -8) as seeds

| **Seed** | **MNI** | | | **k** | **p-FDR** | **Cluster Regions** |
| --- | --- | --- | --- | --- | --- | --- |
|  | **x** | **y** | **z** |  |  |  |
| lNAc | -14 | 8 | -8 | 13248 | 0 | bilateral insular cortex |
|  |  |  |  |  |  | bilateral thalamus |
|  |  |  |  |  |  | bilateral putamen |
|  |  |  |  |  |  | brain stem |
|  |  |  |  |  |  | bilateral orbitofrontal cortex |
|  |  |  |  |  |  | bilateral caudate |
|  |  |  |  |  |  | bilateral Heschl's gyrus |
|  |  |  |  |  |  | bilateral planum temporale |
|  |  |  |  |  |  | bialteral central opercular cortex |
|  |  |  |  |  |  | bilateral frontal operculum cortex |
|  |  |  |  |  |  | bilateral amygdala |
|  |  |  |  |  |  | temporal pole (right) |
|  |  |  |  |  |  | bilateral planum polare |
|  |  |  |  |  |  | posterior supramarginal gyrus (left) |
|  |  |  |  |  |  | bilateral nucleus accumbens |
|  |  |  |  |  |  | parietal operculum cortex (left) |
|  |  |  |  |  |  | bilateral hippocampus |
|  |  |  |  |  |  | bilateral posterior parahippocampal gyrus |
|  |  |  |  |  |  | bilateral pallidum |
|  | 2 | 34 | 18 | 7138 | 0 | anterior cingulate gyrus |
|  |  |  |  |  |  | posterior cingulate gyrus |
|  |  |  |  |  |  | bilateral paracingulate gyrus |
|  | -26 | 50 | 16 | 1320 | 0 | frontal pole (left) |
|  |  |  |  |  |  | middle frontal gyrus (left) |
|  | 30 | 46 | 24 | 960 | 0 | frontal pole (right) |
|  | 0 | -54 | -36 | 462 | 0 | vermis VII-IX (left) |
|  |  |  |  |  |  | cerebellum IX (left) |
|  |  |  |  |  |  | cerebellum crus II (left) |
|  | 20 | -68 | -20 | 382 | 0 | cerebellum IV-VI (right) |
|  |  |  |  |  |  | cerebellum crus I (right) |
|  | -12 | -76 | 36 | 295 | 0 | precuneus |
|  |  |  |  |  |  | cuneal cortex (right) |
|  | 34 | -60 | -52 | 161 | 0.001 | cerebellum VIII-IX (right) |
|  | 14 | -84 | -32 | 89 | 0.01 | cerebellum crus II (right) |
|  | 6 | -60 | 12 | 56 | 0.045 | precuneus cortex |
|  |  |  |  |  |  | intracalcarine cortex (right) |
| rNAc | 14 | 8 | -8 | 7313 | 0 | bilateral insular cortex |
|  |  |  |  |  |  | bilateral putamen |
|  |  |  |  |  |  | bilateral thalamus |
|  |  |  |  |  |  | bilateral orbitofrontal cortex |
|  |  |  |  |  |  | bilateral caudate |
|  |  |  |  |  |  | bilateral amygdala |
|  |  |  |  |  |  | bilateral frontal operculum cortex |
|  |  |  |  |  |  | bilateral nucleus accumbens |
|  |  |  |  |  |  | temporal pole (right) |
|  |  |  |  |  |  | brain stem |
|  |  |  |  |  |  | bilateral planum polare |
|  |  |  |  |  |  | hippocampus (right) |
|  |  |  |  |  |  | bilateral Heschl's gyrus |
|  |  |  |  |  |  | subcallosal cortex |
|  |  |  |  |  |  | posterior parahippocampal gyrus (right) |
|  |  |  |  |  |  | bilateral pallidum |
|  | -2 | 30 | 24 | 4799 | 0 | anterior cingulate gyrus |
|  |  |  |  |  |  | bilateral paracingulate gyrus |
|  |  |  |  |  |  | posterior cingulate gyrus |
|  |  |  |  |  |  | bilateral frontal pole |
|  |  |  |  |  |  | superior frontal gyrus (right) |
|  |  |  |  |  |  | bilateral juxtapositional lobule cortex |
|  |  |  |  |  |  | medial prefrontal cortex |
|  | 32 | 48 | 26 | 330 | 0 | frontal pole (right) |
|  | -24 | 42 | 32 | 269 | 0 | frontal pole (left) |
|  |  |  |  |  |  | middle frontal gyrus (left) |
|  | -18 | -22 | -12 | 61 | 0.047 | hippocampus (left) |
|  | -10 | -40 | -2 | 60 | 0.047 | posterior parahippocampal gyrus (left) |
|  |  |  |  |  |  | posterior cingulate gyrus |
| **Abbreviations:** Montreal Neurological Institute (MNI), left nucleus accumbens (lNAc), right nucleus accumbens (rNAc), false discovery rate (FDR) | | | | | | |

*Table S2.* Associations between adherence-associated rNAC-vmPFC connectivity, headache pain intensity and impact over the course of MBSR+

|  | **Headache Pain Intensity** | | | **Headache Impact** | | |
| --- | --- | --- | --- | --- | --- | --- |
| *Predictors* | *Estimates* | *SE* | *p* | *Estimates* | *SE* | *p* |
| Intercept | 4.48 | 0.28 | **<0.001** | 60.33 | 1.24 | **<0.001** |
| Mid-Treatment | -0.16 | 0.24 | 0.50 | -4.31 | 1.22 | **<0.001** |
| rNAc-vmPFC | 1.12 | 2.38 | 0.64 | -4.01 | 10.68 | 0.71 |
| Post-Treatment | -0.27 | 0.23 | 0.25 | -4.50 | 1.22 | **<0.001** |
| 1-year Follow-Up | -0.08 | 0.24 | 0.74 | -4.04 | 1.23 | **<0.001** |
| Mid-Treatment X rNAc-vmPFC | -0.33 | 2.02 | 0.87 | 14.29 | 10.54 | 0.17 |
| Post-Treatment X rNAc-vmPFC | 2.39 | 2.00 | 0.23 | -5.41 | 10.54 | 0.61 |
| 1-Year Follow-up X rNAc-vmPFC | 1.16 | 2.00 | 0.56 | 11.04 | 10.79 | 0.31 |
| **Random Effects** | | | | | | |
| σ^2^ | 0.73 | | | 20.31 | | |
| τ_00_ | 1.34 _id_ | | | 21.42 _id_ | | |
| N | 43 _id_ | | | 43 _id_ | | |
| Observations | 167 | | | 169 | | |
| Marginal R^2^ / Conditional R^2^ | 0.023 / 0.656 | | | 0.083 / 0.554 | | |

*Notes.* SE = Standard Error. rNAc-vmPFC = functional connectivity between the right nucleus accumbens and ventromedial prefrontal cortex. Follow-up time variables (i.e., Mid-Treatment, Post-Treatment and 1-year Follow-Up) were entered as dummy codes, with baseline as the reference group. σ^2^ = L1 variance; τ_00_ = intercept variance.

*Table S3.* Relationship between meditation practice duration and change in clinical pain over the course of MBSR+

|  | **Headache Frequency** | | | **Headache Impact** | | | **Headache Severity** | | |
| --- | --- | --- | --- | --- | --- | --- | --- | --- | --- |
| *Predictors* | *Estimates* | *SE* | *p* | *Estimates* | *std. Error* | *p* | *Estimates* | *SE* | *p* |
| Intercept | 7.25 | 1.01 | **<0.001** | 60.90 | 2.15 | **<0.001** | 4.31 | 0.50 | **<0.001** |
| Mid-Treatment | -0.74 | 1.07 | 0.49 | -5.07 | 2.17 | **0.02** | -0.02 | 0.44 | 0.96 |
| Home Practice | 0.06 | 0.04 | 0.20 | -0.04 | 0.09 | 0.65 | 0.01 | 0.02 | 0.67 |
| Post-Treatment | -1.82 | 1.06 | 0.09 | -4.90 | 2.17 | **0.02** | -0.20 | 0.43 | 0.64 |
| 1-year Follow-Up | -3.54 | 1.06 | **<0.001** | -5.10 | 2.26 | **0.02** | -0.09 | 0.44 | 0.84 |
| Mid-Treatment X Home Practice | -0.07 | 0.05 | 0.12 | 0.09 | 0.09 | 0.36 | -0.00 | 0.02 | 0.90 |
| Post-Treatment X Home Practice | -0.07 | 0.05 | 0.14 | -0.00 | 0.09 | 0.98 | 0.01 | 0.02 | 0.71 |
| 1-Year Follow-up X Home Practice | 0.02 | 0.05 | 0.71 | 0.09 | 0.10 | 0.38 | 0.01 | 0.02 | 0.76 |
| **Random Effects** | | | | | | | | | |
| σ^2^ | 4.86 | | | 19.77 | | | 0.79 | | |
| τ_00_ | 4.04 _id_ | | | 20.14 _id_ | | | 1.37 _id_ | | |
| N | 47 _id_ | | | 47 _id_ | | | 47 _id_ | | |
| Observations | 187 | | | 182 | | | 183 | | |
| Marginal R^2^ / Conditional R^2^ | 0.181 / 0.553 | | | 0.081 / 0.545 | | | 0.008 / 0.636 | | |

*Notes.* SE = Standard Error. Follow-up time variables (i.e., Mid-Treatment, Post-Treatment and 1-year Follow-Up) were entered as dummy codes, with baseline as the reference group. σ^2^ = L1 variance; τ_00_ = intercept variance.

**Figure S1**

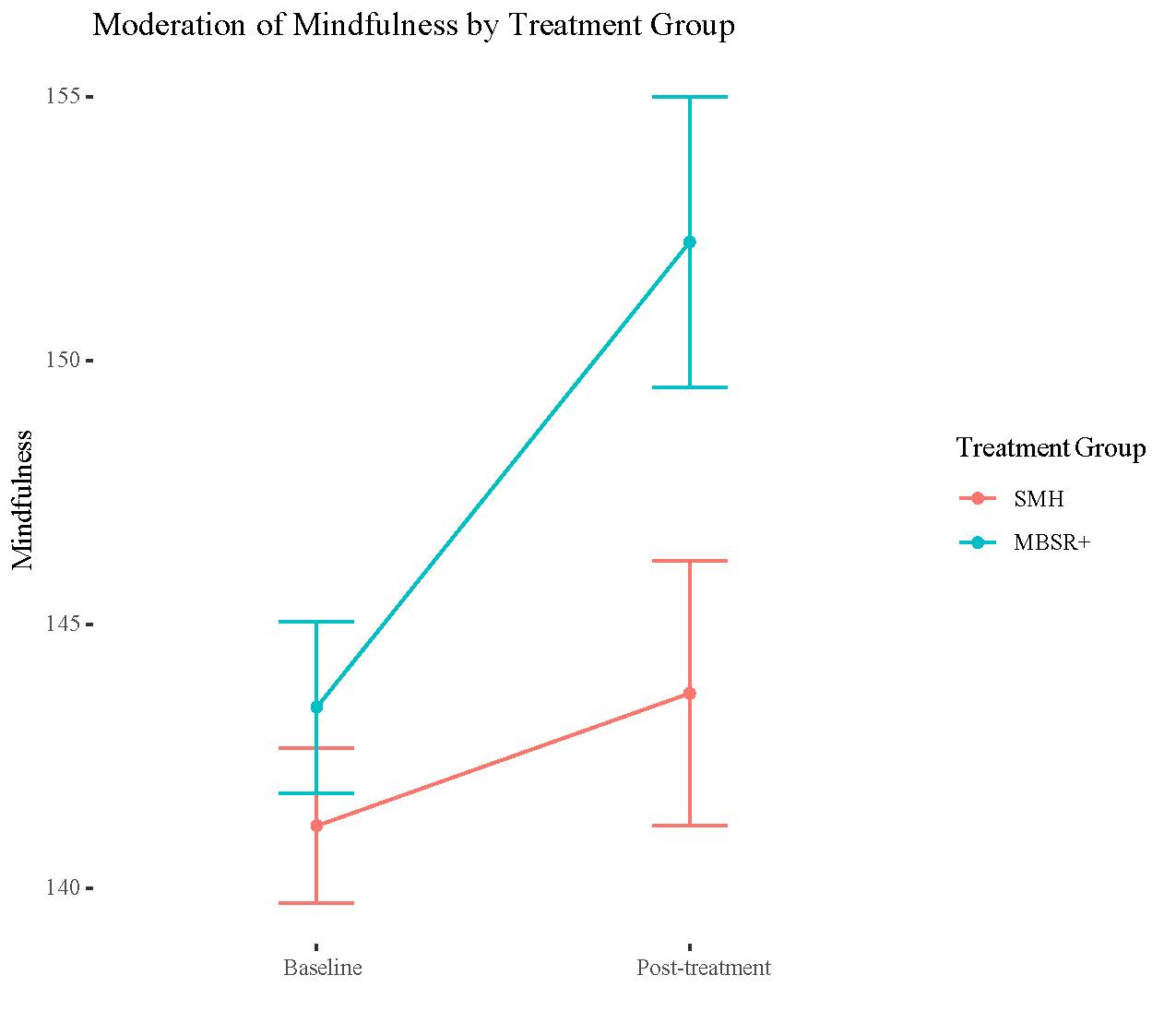

MBSR+ participants demonstrated a significantly greater increase in mindfulness from pre to post-treatment relative to SMH participants. SMH = Stress Management for Headache. MBSR+ = Enhanced Mindfulness-Based Stress Reduction.
